# SARS-CoV-2 Variants of Concern (VOC) Alpha, Beta, Gamma, Delta, and Omicron coincident with consecutive pandemic waves in Pakistan

**DOI:** 10.1101/2022.05.19.22275149

**Authors:** Asghar Nasir, Uzma Bashir Aamir, Akbar Kanji, Azra Samreen, Zeeshan Ansar, Najia K. Ghanchi, Ali Raza Bukhari, Kiran Iqbal Masood, Nazneen Islam, Samina Ghani, M. Asif Syed, Mansoor Wassan, Syed Faisal Mahmood, Zahra Hasan

## Abstract

Identification and monitoring of SARS-CoV-2 Variants of Concern/Interest (VOC/VOIs) is essential to guide public health measures. We report the surveillance of VOCs circulating in Karachi during the pandemic between April 2021 and February 2022. We screened 2150 SARS-CoV-2 PCR positive samples received at the AKUH Clinical Laboratories. VOC was identified using a PCR-based approach targeting lineage-specific mutations using commercially available assays. Of the SARS-CoV-2 PCR positive samples, 81.7% had VOC/VOI, while 18.3% were undetermined. Alpha variants were predominant at 82.5% and 40.3% of the cases in April and May 2021. Beta variants increased in May (29%) and June (42%) and then reduced to 6% by July. Gamma variant cases were at 14.5% and 9% in May and June, respectively. Delta variants first detected in May, increased to comprise 66% of all variants by July, remaining dominant in August, September, October, and November 2021 at 88%, 91%, 91% and 85% respectively. Omicron (BA.1) variants emerged in December, rising to 42% of cases with an increase to 81% by January 2022 and then reducing to 45% in February 2022. Delta variant prevalence was coincident with increased hospital admissions and mortality. The Omicron variant surge was associated with increased daily infections but limited COVID-19 severity. We highlight the predominance of the VOCs identified through a rapid PCR based approach. As this is important to inform a public health response, we propose that a mutation targeted approach can be a rapid, lower cost solution to aid tracking of known VOCs during pandemic waves.

## Introduction

Viral infections are a major global health concern, and new infectious diseases continue to emerge that affect public health. SARS-CoV-2 (previously known as 2019-nCoV), a strain belonging to the coronavirus family that originated in Wuhan, China in December 2019 ^1, 2^has spread to multiple countries around the world and the disease associated with SARS-CoV-2 (COVID-19) is a public health emergency of international concern. According to John Hopkins Coronavirus Resource Center (https://github.com/CSSEGISandData/COVID-19), as of 14^th^ April 2022, SARS-CoV-2 related infections have surpassed over 504 million cases, with 6.19 million deaths globally ^3^.

Five successive COVID-19 waves were documented in Pakistan; March to July 2020, October 2020 - January 2021, April to May 2021^4^ and July to September 2021, and more recently December 2021 to February 2022^5^. To date (14^th^ April 2022), in Pakistan, an estimated 1.53 million COVID-19 cases were reported from 30.3 million SARS-CoV-2 PCR tests, in a population of 220 million people. There have been 30,237 COVID-19 related deaths with a case fatality rate (CFR) of approximately 2 %. Sindh province reported 588,298 COVID-19 cases; more than 50% of these were from the city of Karachi.

Centers for Disease Control and Prevention (https://www.cdc.gov/coronavirus/2019-ncov/variants/variant-info) identified the first four variants of concern (VOC) circulating globally as Alpha (B.1.1.7), Beta (B.1.351), Gamma (P.1) and Delta (B.1.617.1/2) lineages, all characterized by unique signature mutations. VOCs are more transmissible than wild-type SARS-CoV-2 strains, possibly linked to increased disease severity, reduced neutralization by antibodies from previous infection or vaccination resulting in new pandemic surges ^6, 7^. From mid-2021 to November 2021, the Delta variant (originating in India) emerged as the globally dominant variant^8, 9^. In November 2021, a new variant emerged in Botswana, Africa and the World Health Organization (WHO) designated this SARS-CoV-2 variant as Omicron B.1.1.529, as its fifth variant of concern (VOC)^10^. According to a WHO report, the Omicron variant is surpassing the Delta variant as the globally dominant variant and contributing significantly to immune evasion^11^ and antibody evasion^12^.

The gold standard to confirm VOC is through sequencing, however routine genomic surveillance in resource-limited countries like Pakistan is not always readily available. Genomic sequencing is costly and requires complex technical instrumentation and expertise. Coupled with increased global demand and shortage of reagents, sequencing remains a challenge in middle-income countries such as Pakistan. Alternately, PCR-based detection of lineage-specific target mutations can be used to identify known VOCs^13, 14^.

World Health Organization, Pakistan supported the implementation of genomic surveillance for SARS-CoV2 at major diagnostic laboratories including Aga Khan University Hospital (AKUH) as part of a national consortium coordinated by the National Reference Public Health Laboratory at the National Institute of Health, Islamabad.

Sequencing of SARS-CoV-2 strains had identified the introduction of VOC in Pakistan^15-17^; it was not possible to scale up the whole genome sequencing effort country wide to trace and track variants due to resource limitations. Therefore, we applied lineage-specific mutation-specific PCR approach to identify VOC in SARS-CoV-2 PCR positive samples. Sindh province has accounted for 37% of the COVID19 positive cases in Pakistan (https://covid.gov.pk/stats/pakistan). Karachi with 20 million population has been found to comprise 81% of COVID-19 cases across Sindh (https://www.sindhhealth.gov.pk/daily_situation_report). Hence, we focused on samples received at the AKUH Clinical Laboratory from Karachi between April 2021 and February 2022. Using this approach, we were able to identify the emergence of the Alpha, Beta, Gamma, Delta, and more recently of the Omicron variants through the period studied. The objective of this study was to allow for early detection of circulating SARS-CoV2 variants in Pakistan.

## Materials and Methods

This study received approval from the Ethical Review Committee, The Aga Khan University (AKU).

### SARS-CoV-2 PCR testing

Diagnostic testing of respiratory specimens was conducted at the AKU Hospital Clinical Laboratories, Karachi using the COBAS® SARS-CoV-2 assay (COBAS® 6800 Roche platform). AKUH Clinical Laboratories is accredited by the College of American Pathologists, USA.

For this study we recruited samples received at the AKUH Clinical Laboratories from all over Karachi. On each day between April 2021 and February 2022, a retrospective review of positive SARS-CoV-2 specimens from the previous day was conducted. The first 10 positive specimens were identified and collected through consecutive convenience sampling. For VOC testing, RNA was extracted using the QiaAmp RNA MiniKit, Qiagen, according to the manufacturer’s instructions. Briefly, 280 µl NPA was added to 1120 µl AVL buffer without carrier RNA and allowed to incubate for 10 minutes. Next, 560 µl ethanol was added and the mixture was applied to QIAamp Mini column. After spin, the flow through was discarded and washing steps with 500 µl AW1 and 500 µl AW2 buffers were performed. The RNA was eluted in 40 µl AVE buffer and stored at −80°C.

VOC were identified using a PCR based approach targeting lineage specific mutations using commercially available assays; namely NovaType SARS-CoV-2, Gold Standard Diagnostics, Eurofins Technologies, NovaType II SARS-CoV-2, Gold Standard Diagnostics, Eurofins Technologies, Phoenix SARS-CoV-2 Mutant Screen [L452R], Promocure Biotech GmbH, NovaType Select P681R SARS-CoV-2, Gold Standard Diagnostics, Eurofins Technologies, GSD NovaType III SARS-CoV-2, Gold Standard Diagnostics, Eurofins Technologies and TaqPath™ COVID-19 CE-IVD RT-PCR, Applied Bio systems. The specificity target of the probes/primers sequences and their target mutation as provided by the manufacturer is listed Supplementary Table 1-6 of Supplementary File 1.

## Results

The Aga Khan University Hospital Clinical Laboratories has conducted over 530,000 SARS-CoV-2 PCR tests since the start of the pandemic in February 2020 to date in April 2022. The AKUH Clinical Laboratories conducted 238,324 SARS-CoV-2 PCR tests between April 2021 and February 2022, identifying 52,905 positive specimens with an average positivity of 22%, with 17,000 to 32,000 specimens were received per month. The background data set of COVID-19 positive respiratory specimens reported is illustrated in Fig 1A. Our COVID-19 data are in keeping with the national data trends reflecting the first to the fifth waves of the pandemic, Fig. 1B.

**Figure 1.**
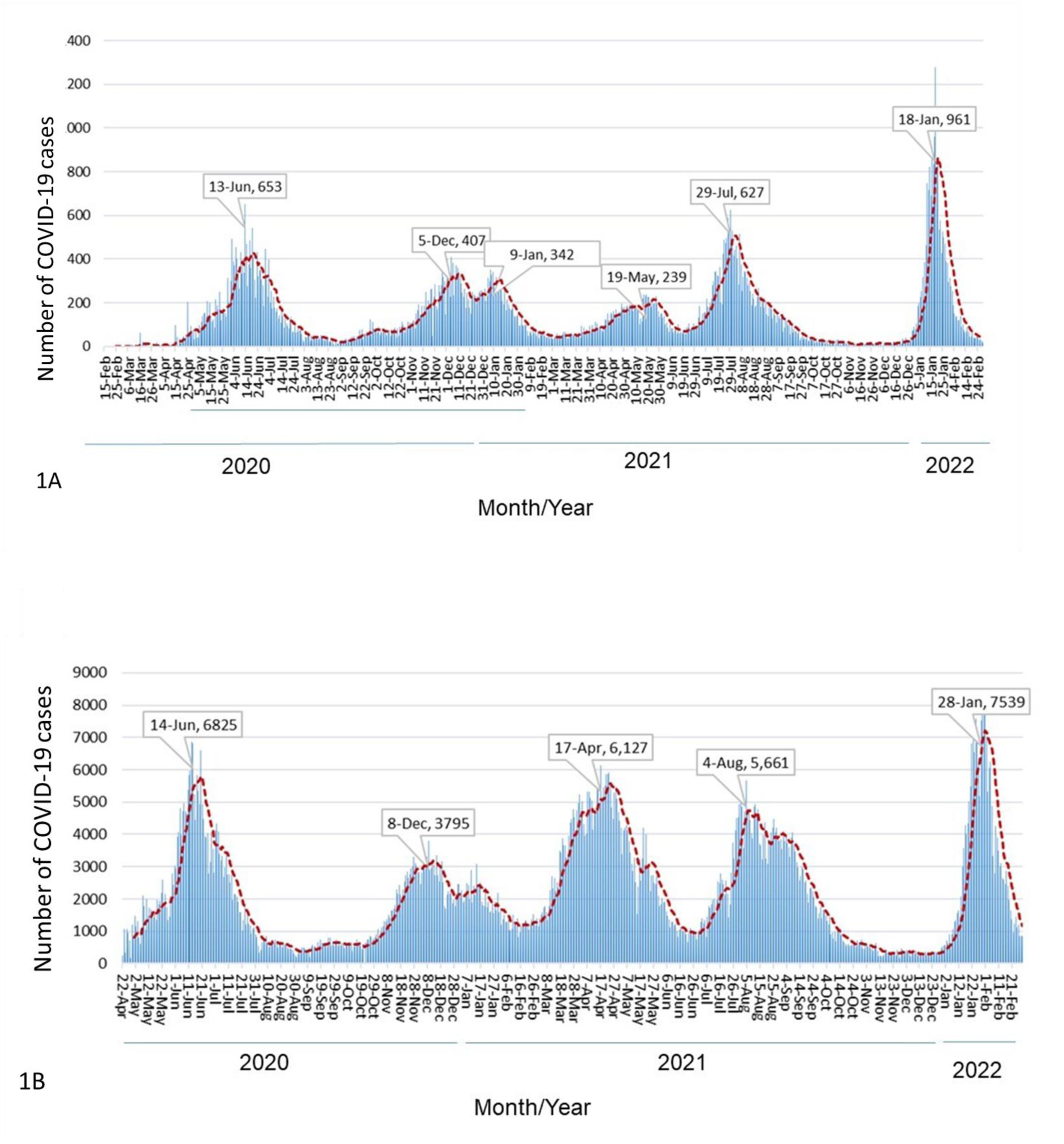
SARS-CoV-2 PCR positive tests reported by the Aga Khan University Hospital Clinical Laboratories, Karachi between February 2020 and February 2022. Figure 1A, the graph depicts the number of SARS-CoV-2 PCR positive respiratory samples reported daily by the AKUH Clinical Laboratories, Karachi, Pakistan. The trend of daily positive cases is depicted with a red dotted trend line. Figure 1B, National data available on the NCOC, government website for the period is depicted.

With an increase in the number of SARS-CoV-2 positive cases at different stages of the pandemic, it was imperative to identify the VOC in the population as a tool for Public Health surveillance. The timeline of the introduction of the assays between April 2021 and February 2022 and the screening workflow is provided in Supplementary Figures 1 and 2 respectively.

Initially, we used the NovaType SARS-CoV-2 assay (April 2021) for the identification of VOC. The assay identified Alpha and Beta variants by detecting the mutations N501Y, A570D. The Alpha variant had both N501Y, A570D mutations whereas the Beta variant only had N501Y mutation only (Supplementary Table S1). NovaType II SARS-CoV-2 assay was used to identify Alpha, Gamma, and Beta variants between May 2021 till December 2021. The assay identified Alpha by detecting N501Y mutation only, Gamma variant by E484K, N501Y mutations and Beta variant by detecting N501Y, E484K, K417N mutations (Supplementary Table S2).

Between May 2021 till August 2021, the Delta variant was identified in parallel with Alpha, Gamma and Beta variants by using the Delta variant-specific assays PhoenixDx SARS-CoV-2 Mutant Screen [L452R] assay and NovaType Select P681R SARS-CoV-2 assay which identified Delta variants by the L452R and P681R mutations respectively (Supplementary Tables 3 and 4). With the introduction and availability of NovaType III SARS-CoV-2 assay in August 2021, it was used for the identification of Delta/Epsilon variant (E484, L452R mutations), Kappa variant (E484Q, L452R mutations) and Beta/Gamma variants (E484K mutation) (Supplementary Table S5).

By December 2021, the emergence of the highly mutated, rapidly spreading Omicron variant had necessitated a change in strategy screening in the wake of soaring Covid-19 case counts and the TaqPath™ COVID-19 CE-IVD RT-PCR was for the identification of the Omicron variant in parallel with the NovaType III assay. Omicron variants resulted in S-gene Target Failure (SGTF) on the TaqPath™ COVID-19 CE-IVD RT-PCR assay (Supplementary Table S6).

Identification of SARS-CoV-2 variant cases was reported on a weekly basis to the Health Department (NIH Situational Reports on the variants identified is available, https://www.nih.org.pk/novel-coranavirus-2019-ncov/), Sindh, Government of Pakistan, National Institutes of Health (Pakistan) and to WHO, Pakistan.

In total, 2150 positive clinical isolates were tested. The highest number of VOC detected in a month was Omicron in January 2022 (297 cases) followed by Delta variant in September 2021 (180 cases), and Beta variant in June 2021 (125 cases) Figure 2A. The prevalence of each VOC/VOI across every month is shown in Figure 2B. Alpha variants were predominant at 82.5%, 40.3% of the cases in April and May 2021. Beta variants increased in May (29%) and June (42%) and then reduced to 6% by July. Gamma variants cases were at 14.5% and 9% in May and June, respectively. Delta variants first detected in May, increased to comprise 66% of all variants by July, remaining dominant in August, September, October, and November 2021 at 88%, 91%, 91% and 85% respectively. Omicron (BA.1) variants emerged in December, rising to 42% of cases with an increase to 81% by January 2022 and then reducing to 45% in February 2022.

**Figure 2.**
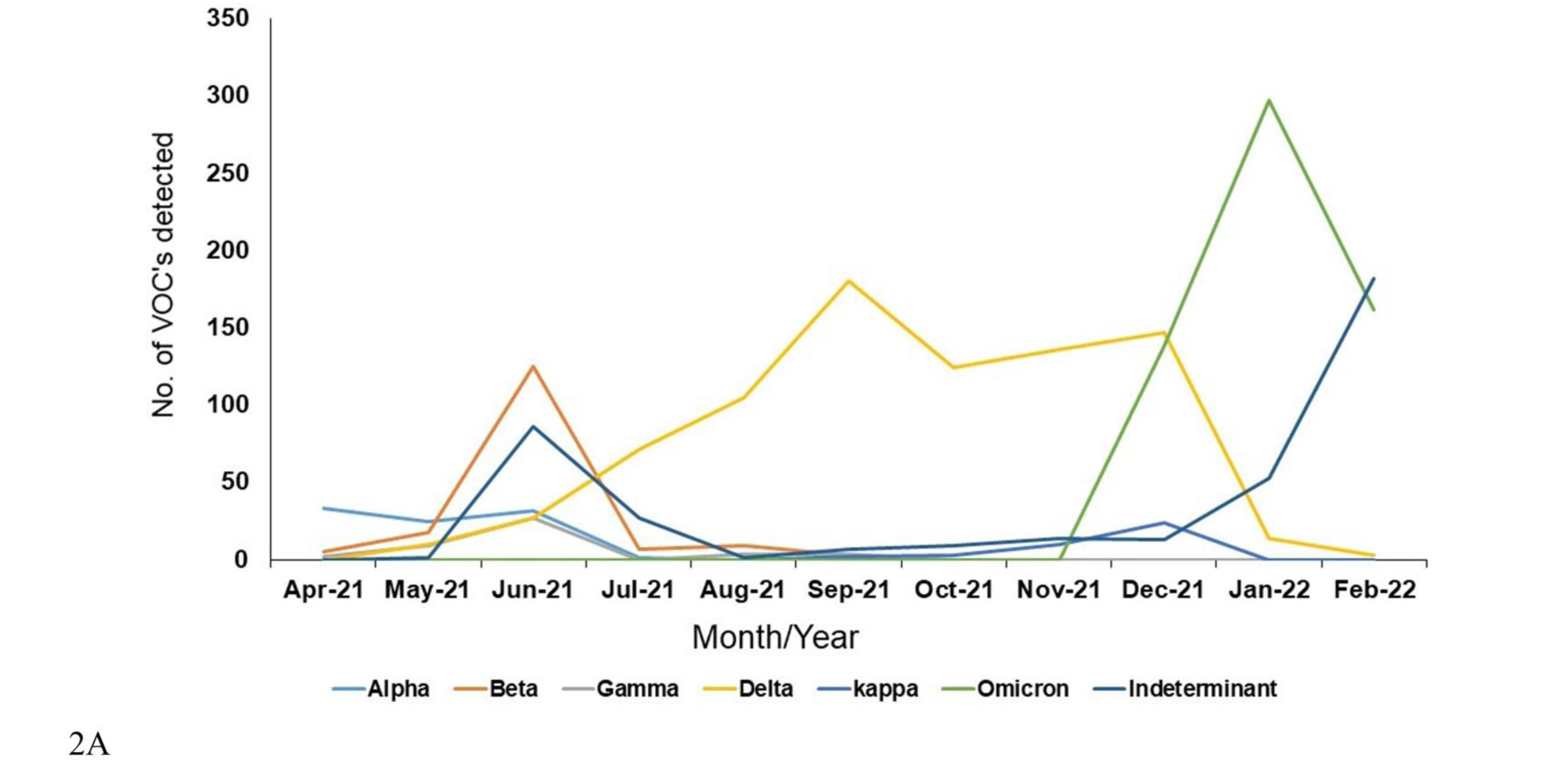
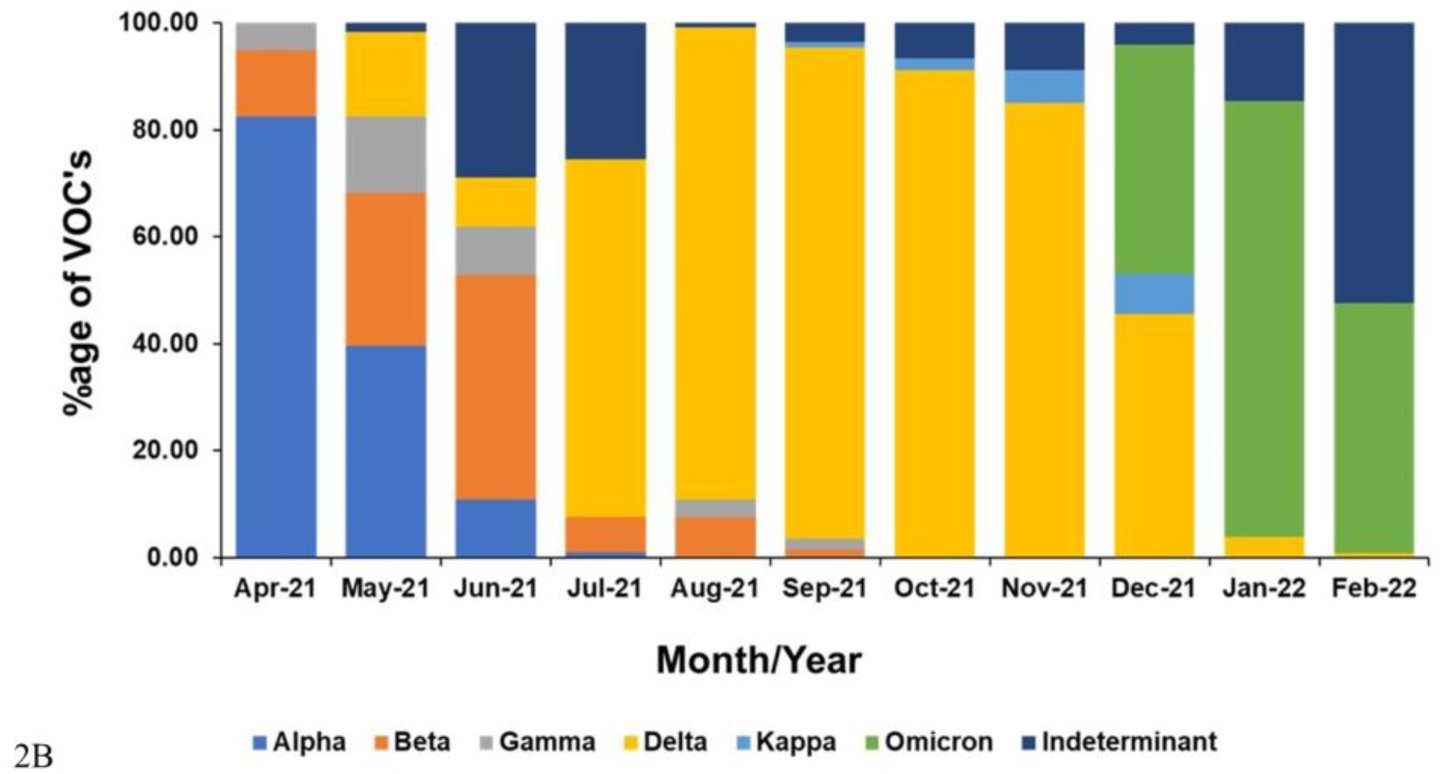
SARS-CoV-2 variants in Karachi. Data presented is of 2150 positive SARS-CoV-2 samples tested Aga Khan University Hospital Clinical Laboratories, Karachi for Alpha, Beta, Gamma, Delta, Kappa and Omicron variants between April 2021 to February 2022 (Figure 2A). Distribution of SARS-CoV-2 variants detected per month is presented Figure 2B.

To determine an association between VOC, waves and COVID-19 associated hospitalization/mortality in Pakistan, we examined data from the COVID-19 Data Repository by the Center for Systems Science and Engineering (CSSE) at Johns Hopkins University. The mortality closely followed the increase in cases during the fourth wave (June 2021 to November 2021). The VOC during this period was the Delta variant. However, in the subsequent, fifth wave January to February where Omicron was predominant, a clear decoupling of the number of cases and the mortality was noted (Supplementary Figure 3 and Supplementary Figure 4).

## Discussion

The AKUH Clinical laboratories received samples for COVID19 PCR testing from all over Karachi and here we focused on those received from Karachi to monitor the spread of VOC through the pandemic.

We report the changing VOC/VOIs associated with waves of the COVID19 pandemic in Pakistan. Our data shows that to date, Pakistan has experienced five different waves of the COVID-19 pandemic. The first wave of COVID-19 peaked in June 2020. The number of positive cases increased again in December 2020, contributing to the second wave of the pandemic in the country. The country’s third wave peaked in May 2021 and was quickly followed up by the fourth wave towards the end of July 2021. The fifth wave of the pandemic peaked in January 2022, and it was associated with the highest number of daily infections since the start of the pandemic.

Between April 2021 till February 2022, we examined 2150 positive cases for VOC/VOIs identification. Of the SARS-CoV-2 PCR positive samples, 81.7% had VOC/VOI while 18.3% were undetermined. Undetermined were identified for further investigation using whole genome sequencing assays (currently underway). Alpha and Beta variants were predominant in April 2021 and May 2021 respectively, coinciding with the third wave of the pandemic in the country. However, by June 2021, the proportion of these variants declined markedly. Between July and November 2021, the Delta variant reached predominance and resulted in the reduction of SARS-CoV-2 variant diversity during the fourth wave of the pandemic. The Omicron variant proportion rapidly increased in December 2021, and it was associated with an increase in daily infections during the Fifth wave of the pandemic. The Delta and Omicron trends observed in Pakistan were no different from those observed in other countries such as the United States and the United Kingdom^18, 19^. Our data is from a single-center however, our COVID-19 data compares well with national trends.

As with other countries^20, 21^, we also note the lower number of deaths, in relation to the cases detected during the fifth wave, when Omicron was the predominant VOC. A decoupling of the number of cases and the mortality during the Omicron surge is documented^22^. This reduced severity is likely a combination of increased pre-existing immunity, vaccine coverage and the inherent properties of the virus. During the previous wave, approximately 30% of the population in Pakistan had received at least a single dose of the vaccine (https://ourworldindata.org/coronavirus/country/pakistan#what-share-of-the-population-has-received-at-least-one-dose-of-the-covid-19-vaccine, Link last accessed 14^th^ April 2022) which had increased to over 50% during the fifth wave. Booster doses had also been introduced, just prior to the fifth wave. On the other hand, the exact contribution of the inherent ability of the Omicron variant to cause severe disease is still under investigation. It is postulated that this may be due to a lack of mutations at the ORF3a gene compared to the previous VOCs^23^, though this is yet to be proven.

Genomic sequencing of SARS-CoV-2 strains can support infection control, epidemiological investigations and viral responses to vaccines and treatments ^24^. Sequencing efforts form Pakistan has contributed to the submission of 2,636 SARS-CoV-2 genomes on GISAID (https://www.gisaid.org/submission-tracker-global/), link last accessed 14 April 2022). Through genomic sequencing, we were one of the first groups in Pakistan to sequence the genomes at the early stages of the pandemic in 2020. In March 2020, strains belonging to L, S, V and GH clades were reported to be circulating in Pakistan but by October 2020, only L and GH strains reported to be circulating ^25^. Our sequencing efforts also identified the introduction of SARS-CoV-2 Alpha variant in Pakistan by international travelers arriving via different flight routes^15^. This genomic surveillance effort was key to implement Trace, Test and Quarantine procedures for arriving travelers by the Department of Health, Government of Sindh.

However, scaling up of SARS-CoV-2 sequencing is limited by technical and financial capacity of countries. We have demonstrated that a PCR-based targeted approach can be used to monitor VOCs in a low-resource setting. However, this approach is unable to fully characterize multiple genetic variations of SARS-CoV-2 lineages and study viral evolution^26^. Furthermore, this method cannot identify minor and novel variants and several strains were ‘undetermined’. Also, as specimens selected for variant screening had lower Ct values, those with a high Ct or lower viral loads may be missed, resulting in an over-representation of VOC. The sample size of our study was relatively small, but the sampling was consistent across the study period. Therefore, the trends in VOC, we observed were similar to those observed elsewhere in Pakistan^27, 28^ and therefore likely representative across the period

A great advantage of the PCR targeted mutation approach is that it allows for the identification of variants in samples with relatively lower viral loads as compared with sequencing. Whole-genome Sequencing of SARS-CoV-2 requires input RNA from samples of low CT values (high viral loads) preferable CT 30 and lower^29^. Consequently, this would preclude variant analysis of respiratory specimens which have a low viral load. In contrast, PCR based mutation detection can be used in samples with lower viral loads of up to Ct 35 and above.

The data presented here highlights the value of linking lab and epidemiological data to response and was helpful in informing a public health response by the provincial and national health authorities supported by WHO thereby leading to targeted enforcement of COVID-19 SOPs and restrictions such as, quarantine, contact tracing and stricter implementation of infection control practices. Meanwhile, vaccination rates have increased in Pakistan, with 120.5 million individuals (55.6% of the population) fully vaccinated (https://coronavirus.jhu.edu/region/pakistan, Link last accessed, 19 April 2022).

## Conclusions

In the future, once a new or emerging variant has been identified by genomic sequencing, it can be tracked using a SARS-CoV-2 variant, PCR-based targeted. This would allow rapid and low-cost evaluation of its transmission.

## Supporting information

sequences

## Data Availability

All data produced in the present study are available upon reasonable request to the authors.

## Acknowledgements

We thank for the following their support in this surveillance initiative; the AKUH Clinical Laboratory particularly, Naima Maniar, Sohail Baloch. Thanks also to the COVID-19 testing staff at the Section of Molecular Pathology, AKUH.

## Funding support

This work received support through a University Research Council Grant, Aga Khan University, Pakistan and the World Health Organization, Pakistan.

## Declarations

The authors have no conflicts of interest to declare.

---

**Supplementary File 1: SARS-CoV-2 variants testing PCR assays**. SARS-CoV-2 variants were identified with a PCR based approach targeting lineage specific mutations using commercially available assays. Supplementary Tables 1 to 6 present the specificity target of the probes/primers sequences and their target mutation of these assays as provided by the manufacturer.

**Supplementary Table 1:**
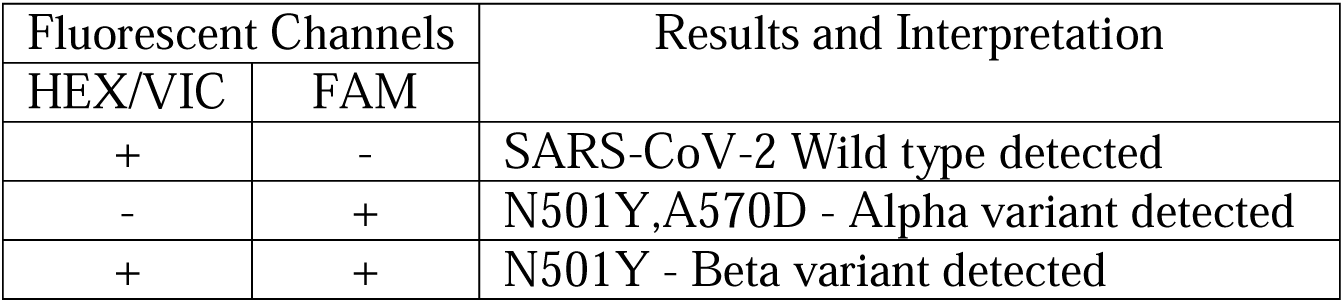
NovaType SARS-CoV-2, Gold Standard Diagnostics, Eurofins Technologies

**Supplementary Table 2:** NovaType II SARS-CoV-2, Gold Standard Diagnostics, Eurofins Technologies

| Fluorescent Channels |  |  |  | Results and Interpretation |
| --- | --- | --- | --- | --- |
| HEX/VIC | CY5 | FAM | ATTO425 |  |
| + | - | - | + | N501, RNase P - Wild type SARS-CoV-2 detected |
| - | - | + | + | N501Y, RNase P - Alpha variant detected |
| + | + | + | + | N501Y, E484K, K417N, RNase P Beta variant detected |
| - | + | + | + | E484K, N501Y, RNase P - Gamma variant detected |
| - | - | - | + | RNase P - Negative - SARS-CoV-2 not detected |

**Supplementary Table 3:** PhoenixDx SARS-CoV-2 Mutant Screen [L452R], Promocure Biotech GmbH

| Fluorescent Channels |  | Results and Interpretation |
| --- | --- | --- |
| FAM | HEX/VIC |  |
| - | - | Negative - SARS-CoV-2 not detected |
| + | - | 452L - Wild type SARS-CoV-2 detected |
| + | + | L452R - Delta variant detected |

**Supplementary Table 4:** NovaType Select P681R SARS-CoV-2, Gold Standard Diagnostics, Eurofins Technologies

| Fluorescent Channels |  |  | Results and Interpretation |
| --- | --- | --- | --- |
| FAM | HEX/VIC | CY5 |  |
| - | - | + | RNase P - Negative SARS-CoV-2 not detected |
| - | + | + | P681 - Wild type SARS-CoV-2 detected |
| + | - | + | 681R, RNase P - Delta variant detected |

**Supplementary Table 5:** GSD NovaType III SARS-CoV-2, Gold Standard Diagnostics, Eurofins Technologies

| Fluorescent Channels |  |  |  |  | Results and Interpretation |
| --- | --- | --- | --- | --- | --- |
| ROX | HEX/VIC | CY5 | FAM | ATTO425 |  |
| - | + | - | - | + | E484, RNase P - Wild type SARS-CoV-2 detected |
| + | + | - | - | + | E484, L452R, RNase P - Delta/Epsilon variant detected |
| + | - | - | + | + | E484Q, L452R, RNase P – Kappa variant detected |
| - | - | + | - | + | E484K, RNase P - Beta/Gamma variant detected |
| - | - | - | - | + | RNase P - Negative - SARS-CoV-2 not detected |

**Supplementary Table 6:**
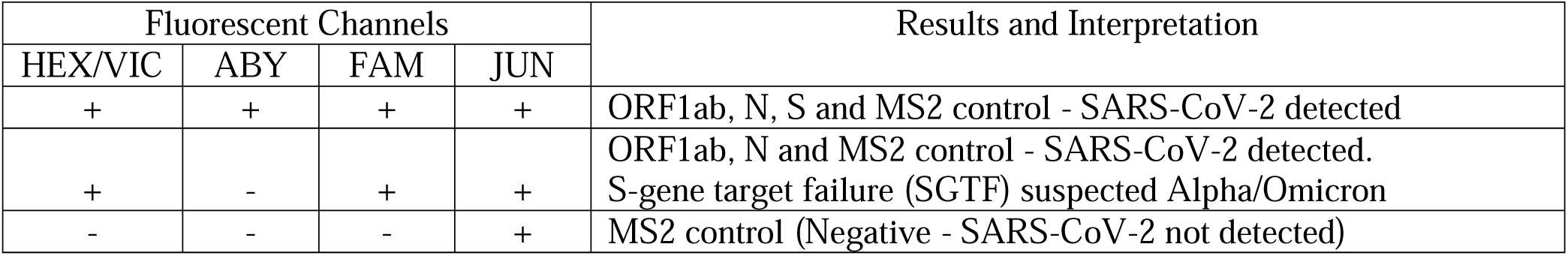
TaqPath™ COVIDJ19 CEJIVD RTJPCR, Applied Bio systems

| Fluorescent Channels |  |  |  | Results and Interpretation |
| --- | --- | --- | --- | --- |
| HEX/VIC | ABY | FAM | JUN |  |
| + | + | + | + | ORF1ab, N, S and MS2 control - SARS-CoV-2 detected |
| + | - | + | + | ORF1ab, N and MS2 control - SARS-CoV-2 detected.<br>S-gene target failure (SGTF) suspected Alpha/Omicron |
| - | - | - | + | MS2 control (Negative - SARS-CoV-2 not detected) |

## Notes

### Competing Interest Statement

The authors have declared no competing interest.

### Funding Statement

This study received support through; University Research Council Grant, Aga Khan University, Pakistan; Rapid Research Grant-236, Higher Education Commission Pakistan and the World Health Organization, Pakistan.

### Author Declarations

This study was approved by the Ethics Review Committee, Aga Khan University.

## References

1. Wang C, Horby PW, Hayden FG, Gao GF. A novel coronavirus outbreak of global health concern. Lancet. 2020;395(10223):470–473.

2. Huang H, Zhang M, Chen C, Zhang H, Wei Y, Tian J, Shang J, Deng Y, Du A, Dai H. Clinical characteristics of COVID-19 in patients with preexisting ILD: A retrospective study in a single center in Wuhan, China. J Med Virol. 2020;92(11):2742–2750.

3. JHU. COVID-19 Data Repository by the Center for Systems Science and Engineering (CSSE) at Johns Hopkins University. 14 April 2022.

4. Dil S, Dil N, Maken ZH. COVID-19 Trends and Forecast in the Eastern Mediterranean Region With a Particular Focus on Pakistan. Cureus. 2020;12(6):e8582.

5. CDC. SARS-CoV-2 Variant Classifications and Definitions. 14 April 2022.

6. Planas D, Veyer D, Baidaliuk A, Staropoli I, Guivel-Benhassine F, Rajah MM, Planchais C, Porrot F, Robillard N, Puech J, Prot M, Gallais F, Gantner P, Velay A, Le Guen J, Kassis-Chikhani N, Edriss D, Belec L, Seve A, Courtellemont L, Pere H, Hocqueloux L, Fafi-Kremer S, Prazuck T, Mouquet H, Bruel T, Simon-Loriere E, Rey FA, Schwartz O. Reduced sensitivity of SARS-CoV-2 variant Delta to antibody neutralization. Nature. 2021;596(7871):276–280.

7. WHO. Tracking SARS-CoV-2 variants. 23rd September, 2021.

8. Thiruvengadam R, Binayke A, Awasthi A. SARS-CoV-2 delta variant: a persistent threat to the effectiveness of vaccines. Lancet Infect Dis. 2022;22(3):301–302.

9. Tian D, Sun Y, Zhou J, Ye Q. The Global Epidemic of the SARS-CoV-2 Delta Variant, Key Spike Mutations and Immune Escape. Front Immunol. 2021;12:751778.

10. Viana R, Moyo S, Amoako DG, Tegally H, Scheepers C, Althaus CL, Anyaneji UJ, Bester PA, Boni MF, Chand M, Choga WT, Colquhoun R, Davids M, Deforche K, Doolabh D, du Plessis L, Engelbrecht S, Everatt J, Giandhari J, Giovanetti M, Hardie D, Hill V, Hsiao NY, Iranzadeh A, Ismail A, Joseph C, Joseph R, Koopile L, Kosakovsky Pond SL, Kraemer MUG, Kuate-Lere L, Laguda-Akingba O, Lesetedi-Mafoko O, Lessells RJ, Lockman S, Lucaci AG, Maharaj A, Mahlangu B, Maponga T, Mahlakwane K, Makatini Z, Marais G, Maruapula D, Masupu K, Matshaba M, Mayaphi S, Mbhele N, Mbulawa MB, Mendes A, Mlisana K, Mnguni A, Mohale T, Moir M, Moruisi K, Mosepele M, Motsatsi G, Motswaledi MS, Mphoyakgosi T, Msomi N, Mwangi PN, Naidoo Y, Ntuli N, Nyaga M, Olubayo L, Pillay S, Radibe B, Ramphal Y, Ramphal U, San JE, Scott L, Shapiro R, Singh L, Smith-Lawrence P, Stevens W, Strydom A, Subramoney K, Tebeila N, Tshiabuila D, Tsui J, van Wyk S, Weaver S, Wibmer CK, Wilkinson E, Wolter N, Zarebski AE, Zuze B, Goedhals D, Preiser W, Treurnicht F, Venter M, Williamson C, Pybus OG, Bhiman J, Glass A, Martin DP, Rambaut A, Gaseitsiwe S, von Gottberg A, de Oliveira T. Rapid epidemic expansion of the SARS-CoV-2 Omicron variant in southern Africa. Nature. 2022;603(7902):679–686.

11. World Health Organization. Enhancing response to Omicron SARS-CoV-2 variant: Technical brief and priority actions for Member States. World Health Organization Headquarters, Geneva, Switzerland. Update,.

12. Iketani S, Liu L, Guo Y, Liu L, Chan JF, Huang Y, Wang M, Luo Y, Yu J, Chu H, Chik KK, Yuen TT, Yin MT, Sobieszczyk ME, Huang Y, Yuen KY, Wang HH, Sheng Z, Ho DD. Antibody evasion properties of SARS-CoV-2 Omicron sublineages. Nature. 2022.

13. Xiong D, Zhang X, Shi M, Wang N, He P, Dong Z, Zhong J, Luo J, Wang Y, Yu J, Wei H. Developing an Amplification Refractory Mutation System-Quantitative Reverse Transcription-PCR Assay for Rapid and Sensitive Screening of SARS-CoV-2 Variants of Concern. Microbiol Spectr. 2022;10(1):e0143821.

14. World Health Organization. Methods for the detection and characterization of SARS-CoV-2 variants: first update, 20 December 2021 (No. WHO/EURO: 2021-2148-41903-62832). World Health Organization. Regional Office for Europe.

15. Nasir A, Bukhari, A.R., Trovão, N.S., Thielen, P.M., Kanji, A., Mahmood, S.F., Ghanchi, N.K., Ansar, Z., Merritt, B., Mehoke, T. and Razzak, S.A. Evolutionary History and Introduction of SARS-CoV-2 Alpha VOC/B. 1.1. 7 in Pakistan Through International Travelers. Virus Evolution. 2022.

16. Umair M, Ikram, A., Rehman, Z., Haider, S.A., Ammar, M., Badar, N., Ali, Q., Rana, M.S. and Salman, M. Genomic surveillance of SARS-CoV-2 reveals emergence of Omicron BA. 2 in Islamabad, Pakistan. medRxiv. 2022.

17. Umair M, Ikram A, Badar N, Haider SA, Rehman Z, Ammar M, Ahad A, Ali Q, Suleman R, Salman M. Tracking down B.1.351 SARS-CoV-2 variant in Pakistan through genomic surveillance. J Med Virol. 2022;94(1):32–34.

18. Lambrou AS, Shirk P, Steele MK, Paul P, Paden CR, Cadwell B, Reese HE, Aoki Y, Hassell N, Zheng XY, Talarico S, Chen JC, Oberste MS, Batra D, McMullan LK, Halpin AL, Galloway SE, MacCannell DR, Kondor R, Barnes J, MacNeil A, Silk BJ, Dugan VG, Scobie HM, Wentworth DE, Caravas J, Kovacs NA, Gerhart JG, Jia Ng H, Beck A, Chau R, Cintron R, Cook PW, Gulvik CA, Howard D, Jang Y, Knipe K, Lacek KA, Moser KA, Paskey AC, Rambo-Martin BL, Nagilla RR, Retchless AC, Schmerer MW, Seby S, Shepard SS, Stanton RA, Stark TJ, Uehara A, Unoarumhi Y, Bentz ML, Burgin A, Burroughs M, Davis ML, Keller MW, Keong LM, L. SS, Lee JS, Madden Jr JC, Nobles S, Owuor DC, Padilla J, Sheth M, Wilson MM, Strain S, Emerging Variants Bioinformatic Working G, Strain S, Emerging Variants NSWG. Genomic Surveillance for SARS-CoV-2 Variants: Predominance of the Delta (B.1.617.2) and Omicron (B.1.1.529) Variants - United States, June 2021-January 2022. MMWR Morb Mortal Wkly Rep. 2022;71(6):206–211.

19. Vihta KD, Pouwels, K.B., Peto, T.E., Pritchard, E., House, T., Studley, R., Rourke, E., Cook, D., Diamond, I., Crook, D. and Matthews, P.C OMICRON-ASSOCIATED CHANGES IN SARS-COV-2 SYMPTOMS IN THE UNITED KINGDOM. medRxiv. 2022.

20. Abdullah F, Myers J, Basu D, Tintinger G, Ueckermann V, Mathebula M, Ramlall R, Spoor S, de Villiers T, Van der Walt Z, Cloete J, Soma-Pillay P, Rheeder P, Paruk F, Engelbrecht A, Lalloo V, Myburg M, Kistan J, van Hougenhouck-Tulleken W, Boswell MT, Gray G, Welch R, Blumberg L, Jassat W. Decreased severity of disease during the first global omicron variant covid-19 outbreak in a large hospital in tshwane, south africa. Int J Infect Dis. 2022;116:38–42.

21. Nyberg T, Ferguson NM, Nash SG, Webster HH, Flaxman S, Andrews N, Hinsley W, Bernal JL, Kall M, Bhatt S, Blomquist P, Zaidi A, Volz E, Aziz NA, Harman K, Funk S, Abbott S, consortium C-GU, Hope R, Charlett A, Chand M, Ghani AC, Seaman SR, Dabrera G, De Angelis D, Presanis AM, Thelwall S. Comparative analysis of the risks of hospitalisation and death associated with SARS-CoV-2 omicron (B.1.1.529) and delta (B.1.617.2) variants in England: a cohort study. Lancet. 2022;399(10332):1303–1312.

22. Madhi SA, Ihekweazu C, Rees H, Pollard AJ. Decoupling of omicron variant infections and severe COVID-19. Lancet. 2022;399(10329):1047–1048.

23. Zhang J, Ejikemeuwa A, Gerzanich V, Nasr M, Tang Q, Simard JM, Zhao RY. Understanding the Role of SARS-CoV-2 ORF3a in Viral Pathogenesis and COVID-19. Front Microbiol. 2022;13:854567.

24. World Health Organization G, Switzerland. Genomic sequencing of SARS-CoV-2: a guide to implementation for maximum impact on public health. 2021.

25. Ghanchi NK, Nasir A, Masood KI, Abidi SH, Mahmood SF, Kanji A, Razzak S, Khan W, Shahid S, Yameen M, Raza A, Ashraf J, Ansar Z, Dharejo MB, Islam N, Hasan Z, Hasan R. Higher entropy observed in SARS-CoV-2 genomes from the first COVID-19 wave in Pakistan. PLoS One. 2021;16(8):e0256451.

26. Boudet A, et al. Limitation of Screening of Different Variants of SARS-CoV-2 by RT-PCR. Diagnostics (Basel). 2021;11(7).

27. Umair M, Ikram, A., Salman, M., Haider, S.A., Badar, N., Rehman, Z., Ammar, M., Rana, M.S. and Ali, Q. Genomic surveillance reveals the detection of SARS-CoV-2 delta, beta, and gamma VOCs during the third wave in Pakistan. Journal of Medical Virology. 2022;94(3):1115–1129.

28. Khan A, Hanif, M., Akhtar Ahmed, S.S., Ghazali, S. and Khanani, R. SARS-CoV-2 UK, South African and Brazilian Variants in Karachi-Pakistan. Frontiers in Molecular Biosciences. 2021.

29. Organization WH. Genomic sequencing of SARS-CoV-2: a guide to implementation for maximum impact on public health, 8 January 2021. 2021.

