## Supplemental Figs for "SARS-CoV-2 Variants of Concern (VOC) Alpha, Beta, Gamma, Delta, and Omicron coincident with consecutive pandemic waves in Pakistan"

**Supplementary Figure 1. The timeline and screening workflow of the PCR assays.** The introduction of the PCR assays between April 2021 and February 2022

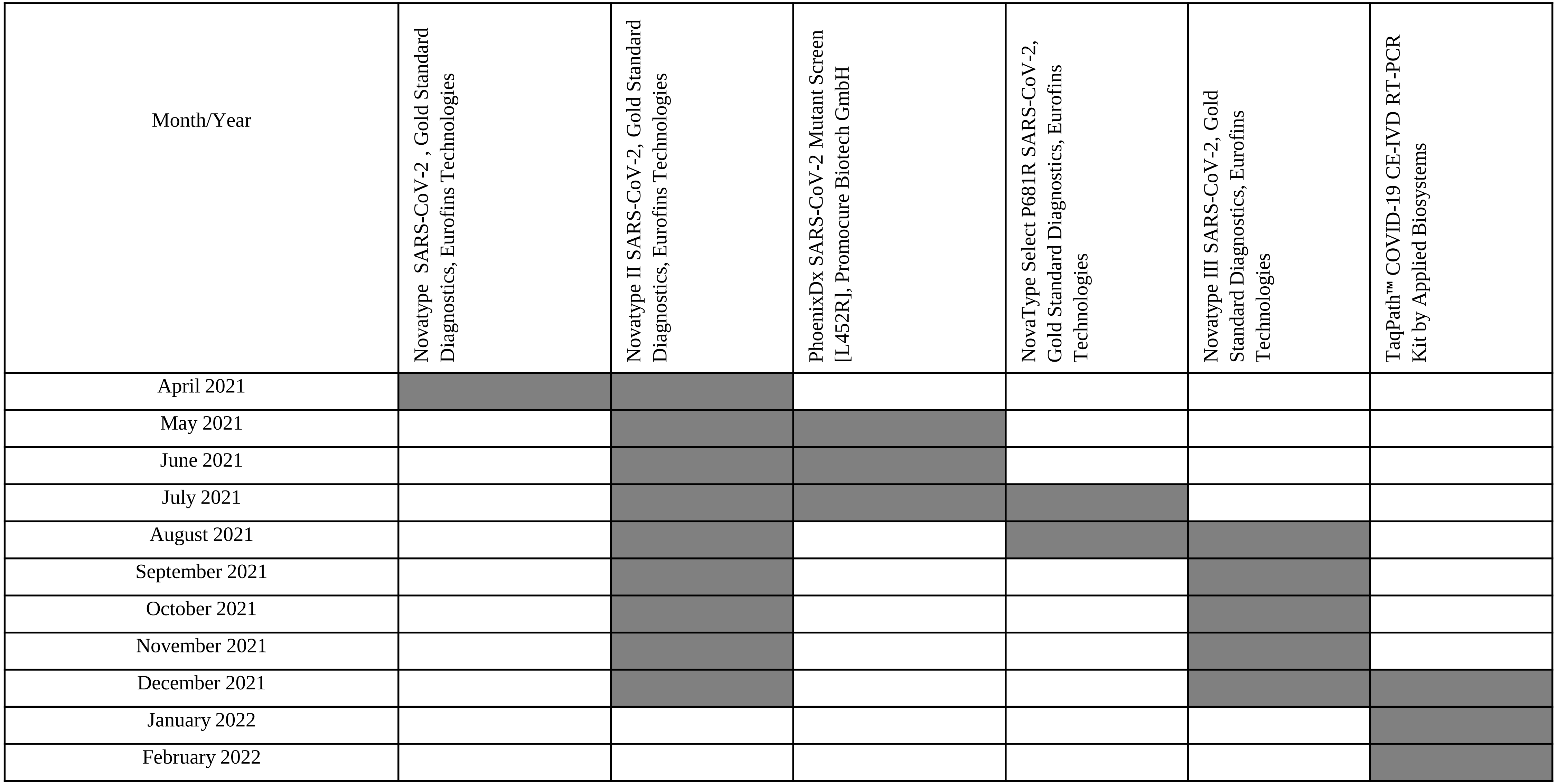

**Supplementary Figure 2**. Workflow of PCR assays utilized

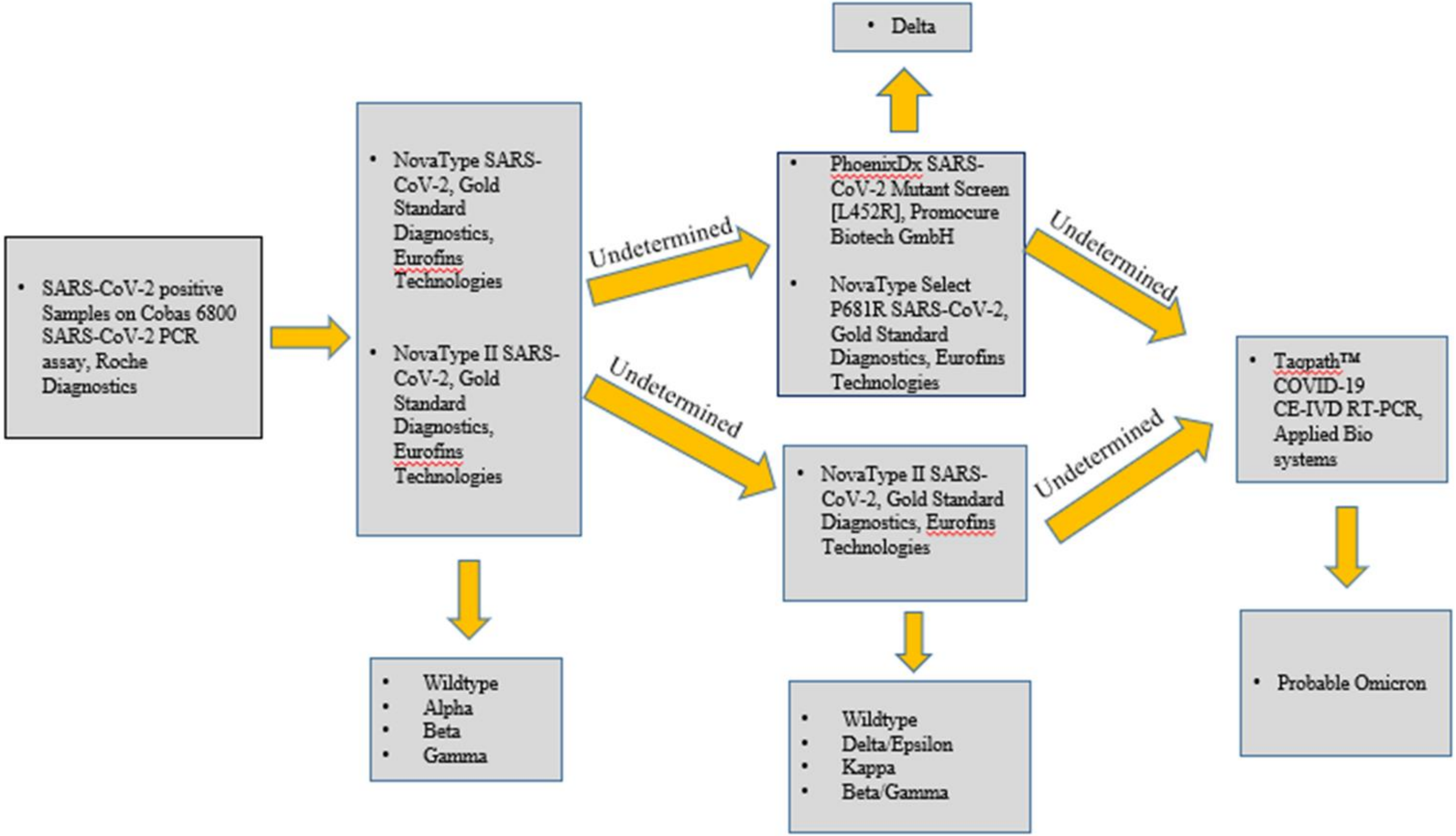

**Supplementary Figure 3. Number of COVID19 positive cases in Pakistan.** Data presented is for the COVID-19 positive cases in Pakistan between months April 2021 till February 2022. Source, John Hopkins, Corona Research Center https://coronavirus.jhu.edu/region/pakistan

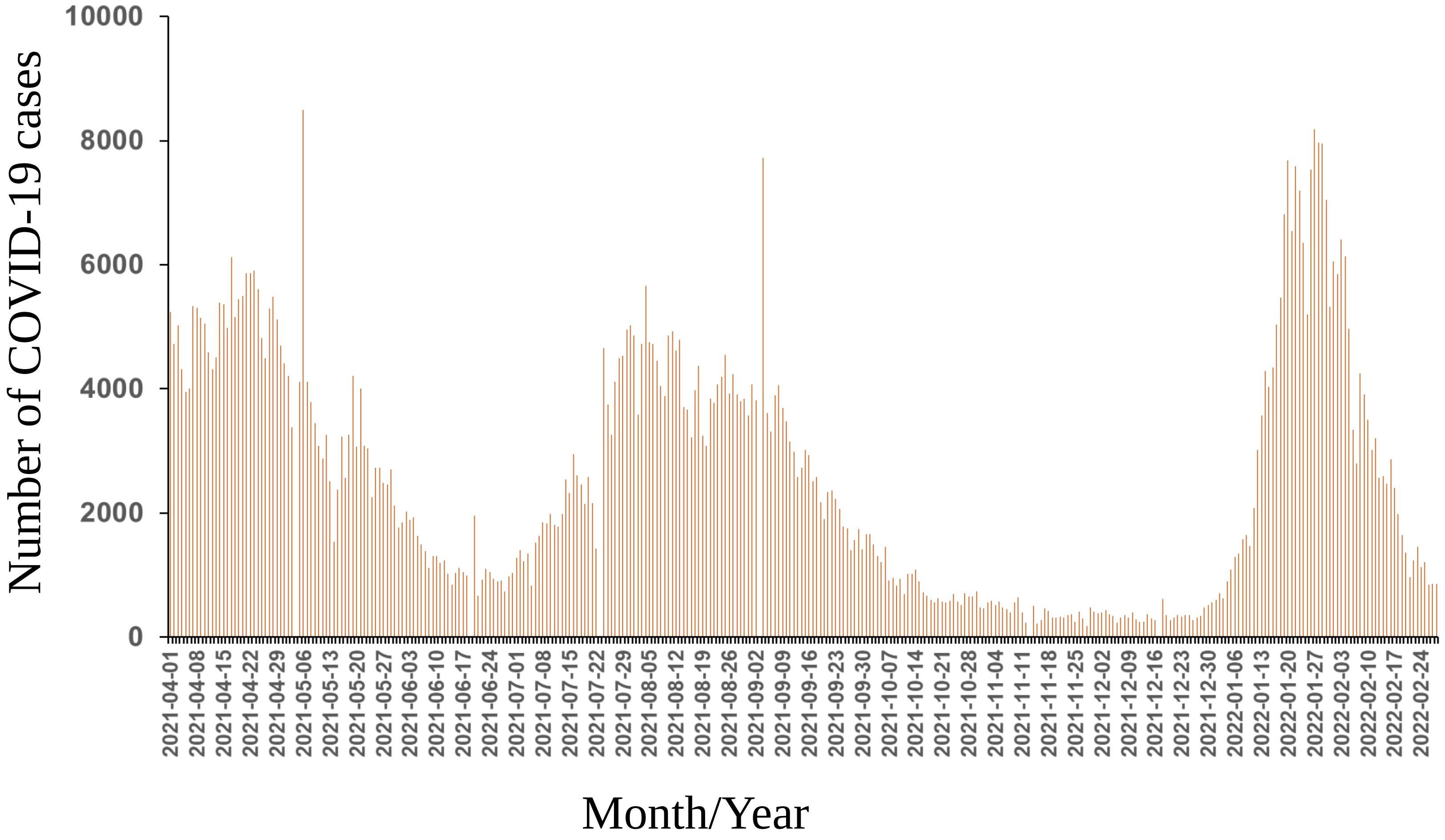

**Supplementary Figure 4. Number of COVID19 related deaths in Pakistan.** Data presented is for the COVID-19 related deaths in Pakistan between months April 2021 till February 2022 Source, John Hopkins, Corona Research Center https://coronavirus.jhu.edu/region/pakistan

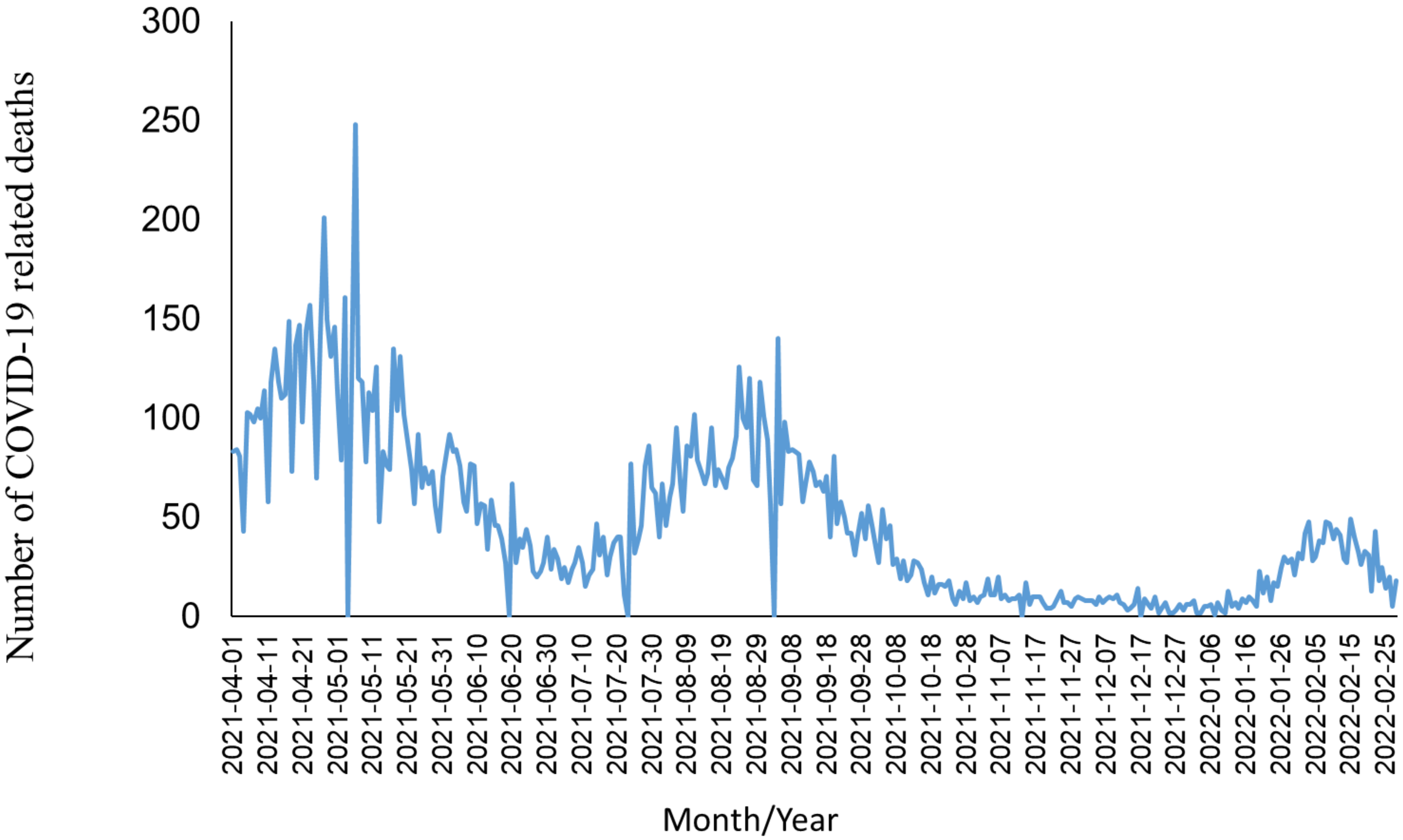
