## Supplementary material for "SARS-CoV-2 Variants of Concern (VOC) Alpha, Beta, Gamma, Delta, and Omicron coincident with consecutive pandemic waves in Pakistan": sequences

| VOC | strain | virus | gisaid_epi_isl | date | region | country |
|---|---|---|---|---|---|---|
| Omicron | hCoV-19/P | betacoronavirus | EPI_ISL_10633680 | Jan-22 | Asia | Pakistan |
| Omicron | hCoV-19/P | betacoronavirus | EPI_ISL_10633681 | Jan-22 | Asia | Pakistan |
| Alpha | hCoV-19/P | betacoronavirus | EPI_ISL_10774011 | Apr-21 | Asia | Pakistan |
| Alpha | hCoV-19/P | betacoronavirus | EPI_ISL_10774012 | Apr-21 | Asia | Pakistan |
| Alpha | hCoV-19/P | betacoronavirus | EPI_ISL_10774013 | May-21 | Asia | Pakistan |
| Beta | hCoV-19/P | betacoronavirus | EPI_ISL_10774014 | May-21 | Asia | Pakistan |
| Beta | hCoV-19/P | betacoronavirus | EPI_ISL_10774015 | May-21 | Asia | Pakistan |
| Omicron | hCoV-19/P | betacoronavirus | EPI_ISL_10774019 | Jan-22 | Asia | Pakistan |
| Alpha | hCoV-19/P | betacoronavirus | EPI_ISL_10813368 | Jan-22 | Asia | Pakistan |
| Delta | hCoV-19/P | betacoronavirus | EPI_ISL_10813369 | Dec-21 | Asia | Pakistan |
| Delta | hCoV-19/P | betacoronavirus | EPI_ISL_11993245 | Jul-21 | Asia | Pakistan |
| Delta | hCoV-19/P | betacoronavirus | EPI_ISL_11993246 | Dec-21 | Asia | Pakistan |
| Beta | hCoV-19/P | betacoronavirus | EPI_ISL_9861904 | Jun-21 | Asia | Pakistan |
| Delta | hCoV-19/P | betacoronavirus | EPI_ISL_9861906 | Jun-21 | Asia | Pakistan |
| Alpha | hCoV-19/P | betacoronavirus | EPI_ISL_9861908 | Jun-21 | Asia | Pakistan |
| Delta | hCoV-19/P | betacoronavirus | EPI_ISL_9861909 | Jun-21 | Asia | Pakistan |
| Alpha | hCoV-19/P | betacoronavirus | EPI_ISL_9861910 | Jun-21 | Asia | Pakistan |
| Beta | hCoV-19/P | betacoronavirus | EPI_ISL_9861911 | Jun-21 | Asia | Pakistan |

| region_exp | country_exp | segment | length | host | age | sex | pangolin_lineage |
|---|---|---|---|---|---|---|---|
| Asia | Pakistan | genome | 29802 | Human | 31-40 | Male | BA.1.1 |
| Asia | Pakistan | genome | 29740 | Human | 21-30 | Male | BA.1.1 |
| Asia | Pakistan | genome | 29818 | Human | 71-80 | Male | B.1.1.7 |
| Asia | Pakistan | genome | 29796 | Human | 71-80 | Male | B.1.1.7 |
| Asia | Pakistan | genome | 29801 | Human | 11-20 | Male | B.1.1.7 |
| Asia | Pakistan | genome | 29821 | Human | >81 | Male | B.1.351 |
| Asia | Pakistan | genome | 29804 | Human | 21-30 | Male | B.1.351 |
| Asia | Pakistan | genome | 29777 | Human | 61-70 | Male | BA.1.1 |
| Asia | Pakistan | genome | 29520 | Human | 21-30 | Female | BA.1.17 |
| Asia | Pakistan | genome | 29473 | Human | 51-60 | Male | B.1.617.2 |
| Asia | Pakistan | genome | 29798 | Human | 61-70 | Male | B.1.617.2 |
| Asia | Pakistan | genome | 29790 | Human | 21-30 | Male | B.1.617.2 |
| Asia | Pakistan | genome | 29736 | Human | 21-30 | Male | B.1.351 |
| Asia | Pakistan | genome | 29750 | Human | 11-20 | Male | B.1.617.2 |
| Asia | Pakistan | genome | 29772 | Human | 21-30 | Male | B.1.1.7 |
| Asia | Pakistan | genome | 29457 | Human | 71-80 | Male | B.1.617.2 |
| Asia | Pakistan | genome | 28956 | Human | 31-40 | Male | B.1.1.7 |
| Asia | Pakistan | genome | 29767 | Human | 21-30 | Female | B.1.351 |

| GISAID_clade | originating_lab | submitting_lab | authors |
|---|---|---|---|
| GRA | Department of Pathol | Department of Pathology | A.R. Bukhari, A. Nasir, A. Ka |
| GRA | Department of Pathol | Department of Pathology | A.R. Bukhari, A. Nasir, A. Ka |
| GR | Department of Pathol | Department of Pathology | A.R. Bukhari, A. Nasir, A. Ka |
| GRY | Department of Pathol | Department of Pathology | A.R. Bukhari, A. Nasir, A. Ka |
| GRY | Department of Pathol | Department of Pathology | A.R. Bukhari, A. Nasir, A. Ka |
| GH | Department of Pathol | Department of Pathology | A.R. Bukhari, A. Nasir, A. Ka |
| GH | Department of Pathol | Department of Pathology | A.R. Bukhari, A. Nasir, A. Ka |
| GRA | Department of Pathol | Department of Pathology | A.R. Bukhari, A. Nasir, A. Ka |
| GRA | Department of Pathol | Department of Pathology | A.R. Bukhari, A. Nasir, A. Ka |
| GK | Department of Pathol | Department of Pathology | A.R. Bukhari, A. Nasir, A. Ka |
| GK | Department of Pathol | Department of Pathology | A. Kanji, A.R. Bukhari, A. Na |
| GK | Department of Pathol | Department of Pathology | A. Kanji, A.R. Bukhari, A. Na |
| GH | Department of Pathol | Department of Pathology | A.R. Bukhari, A. Nasir, A. Ka |
| GK | Department of Pathol | Department of Pathology | A.R. Bukhari, A. Nasir, A. Ka |
| G | Department of Pathol | Department of Pathology | A.R. Bukhari, A. Nasir, A. Ka |
| GK | Department of Pathol | Department of Pathology | A.R. Bukhari, A. Nasir, A. Ka |
| G | Department of Pathol | Department of Pathology | A.R. Bukhari, A. Nasir, A. Ka |
| GH | Department of Pathol | Department of Pathology | A.R. Bukhari, A. Nasir, A. Ka |

| url | date_submitted |
| --- | --- |
| https://www.gisaid.org/ | Mar-22 |
| https://www.gisaid.org/ | Mar-22 |
| https://www.gisaid.org/ | Mar-22 |
| https://www.gisaid.org/ | Mar-22 |
| https://www.gisaid.org/ | Mar-22 |
| https://www.gisaid.org/ | Mar-22 |
| https://www.gisaid.org/ | Mar-22 |
| https://www.gisaid.org/ | Mar-22 |
| https://www.gisaid.org/ | Mar-22 |
| https://www.gisaid.org/ | Mar-22 |
| https://www.gisaid.org/ | Apr-22 |
| https://www.gisaid.org/ | Apr-22 |
| https://www.gisaid.org/ | Feb-22 |
| https://www.gisaid.org/ | Feb-22 |
| https://www.gisaid.org/ | Feb-22 |
| https://www.gisaid.org/ | Feb-22 |
| https://www.gisaid.org/ | Feb-22 |
| https://www.gisaid.org/ | Feb-22 |
